# Ethnic inequalities in physico-chemical, physical and social neighborhood exposures: An individual-level data analysis of 13,926,781 adults

**DOI:** 10.1101/2025.03.18.25321852

**Authors:** Mirthe Muilwijk, Femke Rutters, Jeroen Lakerveld, Petra Elders, Marieke Blom, Karien Stronks, Ilonca Vaartjes, Joline WJ Beulens

**Author notes:** Corresponding author: Dr. Mirthe Muilwijk.

## Abstract

**Highlights:**

– Nation-wide study with data for nearly all adult Dutch inhabitants.
– Ethnic minorities face higher physico-chemical exposures than Dutch-origin inhabitants.
– Food & physical activity environments better for ethnic minorities than Dutch-origin.
– Socio-economic characteristics less favorable for ethnic minorities than Dutch-origin.

**Introduction:** Ethnic minority populations may be disproportionally affected by unhealthy environmental exposures, increasing health inequities. This study aims to identify whether residential neighborhood exposures differ between ethnic groups in the Netherlands.

**Methods:** This cross-sectional study included all adult residents of the Netherlands registered in the national population register on 01/01/2022 (N=13,926,871). Exposure data (physico-chemical, food and physical environment, socio-economic characteristics, health and social well-being) were obtained from Statistics Netherlands, GECCO and the Dutch Health Monitor, and linked to individuals based on geocoded home addresses. Ethnicity was based on country of birth of individuals and their parents. Estimated marginal means were calculated and ethnic differences in exposure determined using multiple linear and logistic regression, adjusted for age and sex, stratified by socio-economic status (SES) and population density.

**Results:** Compared to Dutch-origin, ethnic minority populations had less favorable physico-chemical exposures (e.g. 0.87µg/m^3^ [95%-CI: 0.86;0.88] higher PM2.5 exposure for Moroccans in “high SES-high population density”). Conversely, the food and physical activity environment was more favorable for ethnic minorities (e.g. 1.82km/ha [95%-CI 1.80;1.83] higher bike path density among Turks in the “low SES-low population” density category). Socio-economic characteristics of the environment were generally less favorable for ethnic minorities (E.g. difference between Dutch Caribbeans and Dutch-origin −4.23% [95%-CI −4.35;-4.11] in “high income-high population density”. Ethnic differences in health and social well-being varied. Neighborhood-level smoking was most prevalent among ethnic minorities, while excessive drinking was most prevalent among Dutch-origin. Exposure to vandalism and (sexual)violence was lowest among Dutch-origin and highest among Dutch Caribbean, Moroccans, Turks and Surinamese.

**Conclusion:** Physico-chemical exposure, socio-economic characteristics of the environment and safety from crime were less favorable among ethnic minority populations compared to Dutch-origin. The food and physical activity environment was more favorable for ethnic minorities. Ethnic inequalities were most pronounced among Moroccans, Turks, Surinamese and Dutch Caribbeans compared to Dutch-origin.

## 1. Introduction

Environmental exposures are important upstream drivers of behaviors and health outcomes (Dendup et al. 2018; Lakerveld and Mackenbach 2017; Raphael et al. 2003). Physico-chemical factors such as air quality, noise and temperature can directly impact health. For instance, particulate air pollution exacerbates heart and lung disease, increases all-cause mortality and leads to higher hospital admissions (Pope et al. 1995). Conversely, the food and physical activity environment influence health more indirectly by affecting health-related behaviors and mental health. Greenspace, for example, can enhance the attractiveness of physical activity and improve mental health by providing pleasant scenery and opportunities for social interaction (Chen et al. 2021).

Also social environmental characteristics shape the opportunities and constraints individuals face in maintaining their health (World Health Organization 2011). Residents of neighborhoods with lower socioeconomic positions often experience limited access to healthy food and higher levels of stress, contributing to unhealthy behaviors such as poor dietary choices and tobacco use. The prevalence of health-related measures within communities, such as overweight, can further reinforce such behaviors, as individuals tend to adopt the prevalent habits and attitudes of those around them, also known as social contagion (Powell et al. 2015). A recently published systematic review and meta-analysis highlighted the associations between dimensions of the social environment and cardiometabolic risk factors, underscoring the impact of social disadvantage on conditions such as heart disease and diabetes (Abreu et al. 2024).

Physico-chemical, physical and social neighborhood exposures may contribute to health inequalities, such as between ethnic groups, in case of an unequal distribution. In other words, the differential exposure to environmental factors may partly explain existing health disparities between ethnic groups, such as the observed higher risk for type 2 diabetes and all-cause mortality amongst ethnic minority populations (LaVeist et al. 2009). The environmental justice movement, launched in 1980, has shown that minority and low-income populations are disproportionally exposed to environmental hazards (Evans and Kantrowitz 2002).

However, existing studies on ethnic inequalities in environmental exposures have predominantly focused on air pollution (Ash and Boyce 2018; Liu et al. 2021; van den Brekel et al. 2024), with most research conducted in the United States (Mustansar et al. 2025). Studies have shown that ethnic minority populations in cities in England and the Netherlands experience higher levels of neighborhood air pollution than the host population, even after adjustment for socio-economic position (Fecht et al. 2015). This aligns with recent findings in the Netherlands, indicating that ethnic minority populations are consistently exposed to higher levels of air pollutants, including PM10, PM2.5, NO2, and elemental carbon (EC), compared to the Dutch-origin population (van den Brekel et al. 2024). Although a few smaller studies have examined other environmental characteristics such as noise, the food environment or walkability (Dreger et al. 2019; Jiang et al. 2023; King and Clarke 2015; Poelman et al. 2021), a comprehensive overview of ethnic differences in different environmental domains is currently lacking.

The present study addresses a critical gap by examining a broad range of environmental exposures beyond air pollution, hitherto underexplored, particularly in a European context. Focusing on the Netherlands, this study aims to assess whether and to what extent both physical and social health-related neighborhood exposures differ between ethnic groups. We consider the complex intertwinement between ethnicity, socio-economic status (SES) and population density, building on evidence from European studies that suggest heterogeneous associations between SES and environmental exposure depending on the type of exposure and regional context (Morrens et al. 2012; Saucy et al. 2024).

## 2. Methods

### 2.1 Population and study design

This cross-sectional study utilized data from all adult residents (≥18 years) of the Netherlands registered in the national population register maintained by Statistics Netherlands as of January 1, 2022. The dataset included individuals with complete information on address, ethnicity, age, sex, income and population density, ensuring high coverage due to mandatory registration. The most recent environmental data were extracted from Statistics Netherlands, the Geoscience and Health Cohort Consortium (GECCO) (Lakerveld et al. 2020; Timmermans et al. 2018a) and the Dutch Health Monitor as of January 1, 2022. These data were linked to individuals based on their geocoded home addresses. GECCO provides longitudinal environmental data at various geospatial levels in the Netherlands. Address-level exposure data obtained from GECCO were aggregated to mean values of 6-digit postal code areas, representing small areas that can be crossed in about 8 minutes on foot. The health monitor periodically maps the health, well-being and lifestyle of the Dutch population, with data collected by Statistics Netherlands in collaboration with municipal health services (GGD-en) and the National Institute for Public Health and the Environment (RIVM) using online and paper questionnaires. A stratified sampling method ensures a representative sample of residents per neighborhood across the Netherlands. This study is based on secondary data analyses of existing datasets. All data were pseudonymized prior to access, and no directly identifiable personal information was available to the researchers. The Medical Ethical Committee of Amsterdam UMC (correspondence 2025.0206) declared that this study does not fall under the scope of the Dutch Medical Research Involving Human Subjects Act (WMO), since persons in the study were not subjected to any actions and no particular behavior was imposed. Access to the data was granted following the standard procedures and data protection regulations of the respective data providers. This study was conducted in accordance with the General Data Protection Regulation (GDPR) and relevant national privacy regulations.

### 2.2 Ethnicity

Ethnicity was determined according to the Statistics Netherlands’ definition for migration background, based on the country of birth of individuals and their parents (Centraal Bureau voor de Statistiek 2021). For first-generation individuals (born outside the Netherlands), the ethnic group was determined by the person’s country of birth. For second-generation individuals (born in the Netherlands with one or both parents born abroad), the ethnic group was based on the parents’ country of birth or, in the case of different parents’ countries of birth, on the mother’s country of birth. Country of birth has proven to be a valid indicator for differentiating between ethnic populations in the Netherlands (Stronks et al. 2009). Study outcomes were stratified by common immigration countries (Morocco, Türkiye, Suriname, Dutch Caribbean, Indonesia), Europe (excluding the Netherlands), and non-European (excluding common immigration countries), in line with the ‘new classification of population by origin’ defined by Statistics Netherlands (Centraal Bureau voor de Statistiek 2021). The terms Moroccans, Turks, Surinamese, Dutch Caribbean, Indonesian, European, and non-European refer to individuals with a non-Dutch background from these respective regions or countries. “Dutch” refers here to individuals who were born in the Netherlands and whose parents were also born in the Netherlands.

### 2.3 Exposure data

We assessed four key domains of neighborhood exposures: physico-chemical, food and physical activity environment, socio-economic characteristics, and health and social well-being. Physico-chemical exposures included air pollution, noise and temperature. Food and physical activity environment was evaluated based on access to healthy food options and opportunities for physical activity. Socio-economic characteristics encompassed factors such as liveability scores, election turnout, labor participation and rates of living below the social minimum. Health and social well-being included prevalence of excessive drinking, smoking and safety from crime. An overview of exposure data and the measurement methods are shown in Table 1.

**Table 1:** Overview of the included exposure variables.

| Domain | Exposure | Year | Level | Description |
| --- | --- | --- | --- | --- |
| Physico-chemical | Air pollution | 2021 | PC6 | Metrics include PM10, PM2.5, NO2, and EC, Average concentrations modeled using Land Use Regression (LUR) models at 25x25m resolution (Eeftens et al. 2012). |
| Physico-chemical | Noise | 2016-2020 | PC6 | Cumulative yearly average noise exposure from roads, rail, air, industry and wind turbines. Modeled with the Empara noise tool at 25x25m resolution. Measured in Level day-evening-night (Lden) and expressed in A-weighted decibels (dB(A)). |
| Physico-chemical | Temperature | 2021 | PC6 | Data from KNMI weather stations interpolated to 25x25m grids. Average lowest daily temperature over summer (jun-aug) calculated. Urban heat island (UHI) effect corrected using RIVM UHI dataset. |
| Food and physical activity environment | Food Healthiness Index | 2020 | PC6 | Calculated using LOCATUS data. Kernel density Healthiness Score (KDHS) based on food environment healthiness index (FEHI) for each retailer in the neighborhood (Timmermans et al. 2018b). |
| Food and physical activity environment | Walkability index | 2017-2019 | PC6 | Composite score based on seven components: population density, retail and service destination density, land-use mix, street connectivity, greenspace, sidewalk surface area density, and public transport density (Lam et al. 2022). |
| Food and physical activity environment | Bicycle path density | 2019 | PC6 | Density of bicycle paths per hectare, based on combined data from Dutch topographical map, large-scale topography basic registry and national cycle platform. |
| Food and physical activity environment | Physical activity resources | 2017 | PC6 | Point density of sport accommodations within a 1000m radius, registered by the Mulier institute. |
| Food and physical activity environment | Greenspace | 2017 | PC6 | Density of greenspace within a 1000m buffer, including high vegetation, shrubs, and vegetation, but excluding grass on lawns, meadows, and agricultural crops. Data from RIVM on a 10x10m resolution, resampled to 25x25m grids. |
| Socio-economic characteristics | Liveability | 2020 | PC6 | Scores based on 45 environment characteristics (Mandemakers et al. 2021). Mapped for PC6 areas. |
| Socio-economic characteristics | Socioeconomic position | 2017 | PC4 | Neighborhood-level scores, based on education level, income, and labor market position (Wagtendonk 2019). |
| Socio-economic characteristics | Voters | 2019 | PC4 | Number of voters according to Open State Foundation were divided by number of voters called in a PC4 area. |
| Socio-economic characteristics | Social minimum | 2022 | Neighborhood | Key figures for districts and neighborhoods 2022 from Statistics Netherlands. Private households where the main breadwinner (or partner) has an income throughout the year and is not dependent on student finance. |
| Socio-economic characteristics | Low educational level | 2022 | Neighborhood | Key figures for districts and neighborhoods 2022 from Statistics Netherlands. Low education level was equivalent to International Standard Classification of Education levels 0-2. |
| Socio-economic characteristics | Labor participation | 2022 | Neighborhood | Key figures for districts and neighborhoods 2022 from Statistics Netherlands. Net labor participation among those aged 15-75 years old. |
| Health and social well-being | Overweight | 2020 | Neighborhood | Data from the health monitor adults and elderly 2020. Calculated for each neighborhood using the Small Area Estimates for Policy makers (SMAP) methodology. |
| Health and social well-being | Smoking | 2020 | Neighborhood | Data from the health monitor adults and elderly 2020. Calculated for each neighborhood using the Small Area Estimates for Policy makers (SMAP) methodology. |
| **Health and social well-being** | **Excessive drinking** | 2020 | Neighborhood | Data from the health monitor adults and elderly 2020. Calculated for each neighborhood using the Small Area Estimates for Policy makers (SMAP) methodology. |
| **Health and social well-being** | **Comply with exercise guideline** | 2020 | Neighborhood | Data from the health monitor adults and elderly 2020. Calculated for each neighborhood using the Small Area Estimates for Policy makers (SMAP) methodology. |
| **Health and social well-being** | **Vandalism** | 2018 | Neighborhood | Data from police registrations 2018, linked to neighborhood codes using center of gravity determinants by Statistics Netherlands. |
| **Health and social well-being** | **Sexual violence** | 2018 | Neighborhood | Data from police registrations 2018, linked to neighborhood codes using center of gravity determinants by Statistics Netherlands. |

### 2.4 Confounders and effect modifiers

Age (in years on December 31, 2021) and birth sex were considered as confounders, and were obtained from the National Population Register. SES and population density were effect modifiers. Individual income for 2022 was used as a proxy for SES at the individual level, defined as the yearly disposable income of the individual’s household, adjusted for household size. Income was categorized into high (≥58 percentile) and low (<58 percentile) based on the median. Population density was sourced from the key figures for districts and neighborhoods provided by Statistics Netherlands. Areas were categorized into low density (<2000 inhabitants/km^2^) or high density (≥2000 inhabitants/km^2^), with the latter representing medium-intensive construction areas (Rijksinstituut voor Volksgezondheid en Milieu: Bilthoven).

### 2.5 Statistical analysis

All analyses were conducted stratified by the effect modifiers SES and population density. Descriptive statistics are shown for baseline characteristics. Estimated marginal means were calculated for each of the physical and social neighborhood exposures. Differences between ethnic groups were assessed using multiple linear regression for continuous variables and logistic regression for categorical variables, with Dutch-origin as the reference category. Non-normally distributed data were ln-transformed before analyses and back-transformed by exponentiation after analysis. Analyses were adjusted for age and sex. All statistical analyses were conducted in R studio v4.2.2 (R Core Team 2021).

### 2.6 Sensitivity analyses

Where relevant, exposure areas for buffers and radii around addresses of 1000m were selected, with additional buffers of 500 and 1500/1650m used in sensitivity analyses. Where available, we explored whether alternative variables would yield different findings. These variables included: households below 110% and 120% of the social minimum, households belonging to the 40% lowest income households, households with a low income, average residence value, voters in 2014, obesity, non-drinkers, minutes of exercise, compliance with bone and muscle strengthening exercises guideline, compliance with balance exercise guideline, thefts from home/sheds/etc. Additionally, we evaluated whether ethnic differences in environmental exposures were consistent for first- and second-generation migrants. Further analyses used 2022 key figures for districts and neighborhoods to assess the percentage of Moroccans, Surinamese, Turks and Dutch Caribbean individuals living in the environment of each ethnic group.

## 3. Results

### 3.1 Study population and baseline characteristics

As of January 1^st^ 2022, the Netherlands had a total population of 17,590,672 inhabitants, of whom 14,289,834 were aged ≥18 years (Supplementary Figure 1). We successfully linked 14,281,869 individuals to a valid address (99.9%). Income data was missing for 2.49% of the populations, with 1.74% due to institutionalization. A total of 0.15% had missing data for population density. Data for age, sex and ethnicity were complete. Ultimately, 13,906,254 individuals (97.4%) were included in the study population. The majority of the included individuals were Dutch-origin (75.9%), followed by European (7.4%), non-European (6.9%), Turks (2.4%), Indonesians (2.3%), Surinamese (2.1%), Moroccans (2.1%), and Dutch Caribbean (0.9%). 40.2% of the individuals fell into the “high SES-high population density” category, followed by the “low SES-high population density” category (39.0%), the “low SES-low population density” category (10.6%), and the “high SES-low population density” category (9.9%). The percentage of women ranged from 45.8% among Turks in the “high SES-high population density” category to 56.8% among Surinamese in the “low SES-low population density” category (Table 2). The median age was lowest among Dutch Caribbean individuals in the “low SES-low density” category (34.0 [IQR 25.0; 51.0] years) and highest among Indonesians in the “low SES-high density” category (60.0 [IQR 47.0; 71.0]).

**Table 2:** Baseline characteristics stratified by ethnicity, socio-economic status and population density.

|  |  | Dutch-origin | Indonesian | Moroccan | Surinamese | Turkish | Dutch Caribbean | European | non-European |
| --- | --- | --- | --- | --- | --- | --- | --- | --- | --- |
| **High SES – Low density** | **N people** | 1,095,729 | 31,022 | 21,491 | 30,897 | 30,148 | 9,515 | 97,464 | 85,713 |
|  | **Sex (N women)** | 517529 (47.2%) | 15413 (49.7%) | 10336 (48.1%) | 15426 (49.9%) | 13867 (46.0%) | 4385 (46.1%) | 49598 (50.9%) | 42791 (49.9%) |
|  | **Age (Median)** | 49.0 [33.0; 60.0] | 54.0 [43.0; 64.0] | 35.0 [26.0; 47.0] | 44.0 [31.0; 56.0] | 37.0 [28.0; 48.0] | 38.0 [29.0; 51.0] | 41.0 [31.0; 54.0] | 36.0 [30.0; 47.0] |
| **High SES – High density** | **N people** | 4,609,045 | 133,685 | 54,378 | 92,891 | 87,026 | 32,097 | 322,719 | 263,912 |
|  | **Sex (N women)** | 2188422 (47.5%) | 66116 (49.5%) | 26199 (48.2%) | 46818 (50.4%) | 39878 (45.8%) | 14908 (46.4%) | 166431 (51.6%) | 134516 (51.0%) |
|  | **Age (Median)** | 50.0 [33.0; 60.0] | 55.0 [45.0; 64.0] | 36.0 [26.0; 48.0] | 44.0 [31.0; 56.0] | 38.0 [28.0; 49.0] | 41.0 [30.0; 53.0] | 44.0 [32.0; 56.0] | 37.0 [30.0; 49.0] |
| **Low SES – Low density** | **N people** | 939,490 | 27,030 | 70,088 | 54,385 | 63,205 | 25,199 | 146,161 | 155,251 |
|  | **Sex (N women)** | 49,9251 (53.1%) | 14,807 (54.8%) | 35,811 (51.1%) | 30,872 (56.8%) | 32,016 (50.7%) | 13,514 (53.6%) | 77,548 (53.1%) | 78,682 (50.7%) |
|  | **Age (Median)** | 56.0 [34.0; 71.0] | 58.0 [43.0; 69.0] | 42.0 [29.0; 54.0] | 48.0 [32.0; 62.0] | 42.0 [30.0; 55.0] | 34.0 [25.0; 51.0] | 39.0 [27.0; 58.0] | 36.0 [27.0; 51.0] |
| **Low SES – High density** | **N people** | 3,905,050 | 122,511 | 147,738 | 119,266 | 152,700 | 61,756 | 457,947 | 460,745 |
|  | **Sex (N women)** | 2109372 (54.0%) | 66817 (54.5%) | 74550 (50.5%) | 66404 (55.7%) | 76507 (50.1%) | 32613 (52.8%) | 245321 (53.6%) | 233971 (50.8%) |
|  | **Age (Median)** | 59.0 [38.0; 73.0] | 60.0 [47.0; 71.0] | 41.0 [29.0; 54.0] | 46.0 [32.0; 61.0] | 41.0 [30.0; 54.0] | 38.0 [27.0; 54.0] | 45.0 [31.0; 66.0] | 38.0 [28.0; 51.0] |

Data are N (numbers) and % (percentages) or median and interquartile ranges. Income was used as proxy for socio-economic status and categorized into high ( $\geq 58$ ) and low ( $< 58$ ) percentiles. Population density was categorized into high ( $\geq 2000$ inhabitants/km<sup>2</sup>) and low ( $< 2000$ inhabitants /km<sup>2</sup>).

### 3.2 Physico-chemical exposures

Exposure to physico-chemical factors, covering elemental carbon, nitrogen dioxide, PM10, PM2.5, noise and temperature, was consistently higher among ethnic minority populations compared to the Dutch-origin population. For example, PM10 exposure was 0.87µg/m^3^ [95%-CI 0.86; 0.88] higher for Moroccans compared to Dutch-origin in the “high SES-high population density” category (Figure 1). The largest ethnic differences were observed among Moroccans, Turks, Surinamese and Dutch Caribbeans, followed by non-Europeans, Europeans and Indonesians. The ethnic differences in exposure were more pronounced in areas with low population density compared to high population density areas. For instance, non-Europeans in the “high SES-high population density” had a 2.15Lden higher exposure to noise compared to Dutch-origin, while this difference was 4.10Lden in the “high SES-low population density” category. While the range of exposure between the lowest and highest exposure strata differed slightly among the Dutch-origin population, the range of exposure was much larger for people from ethnic minority backgrounds.

**Fig 1:**
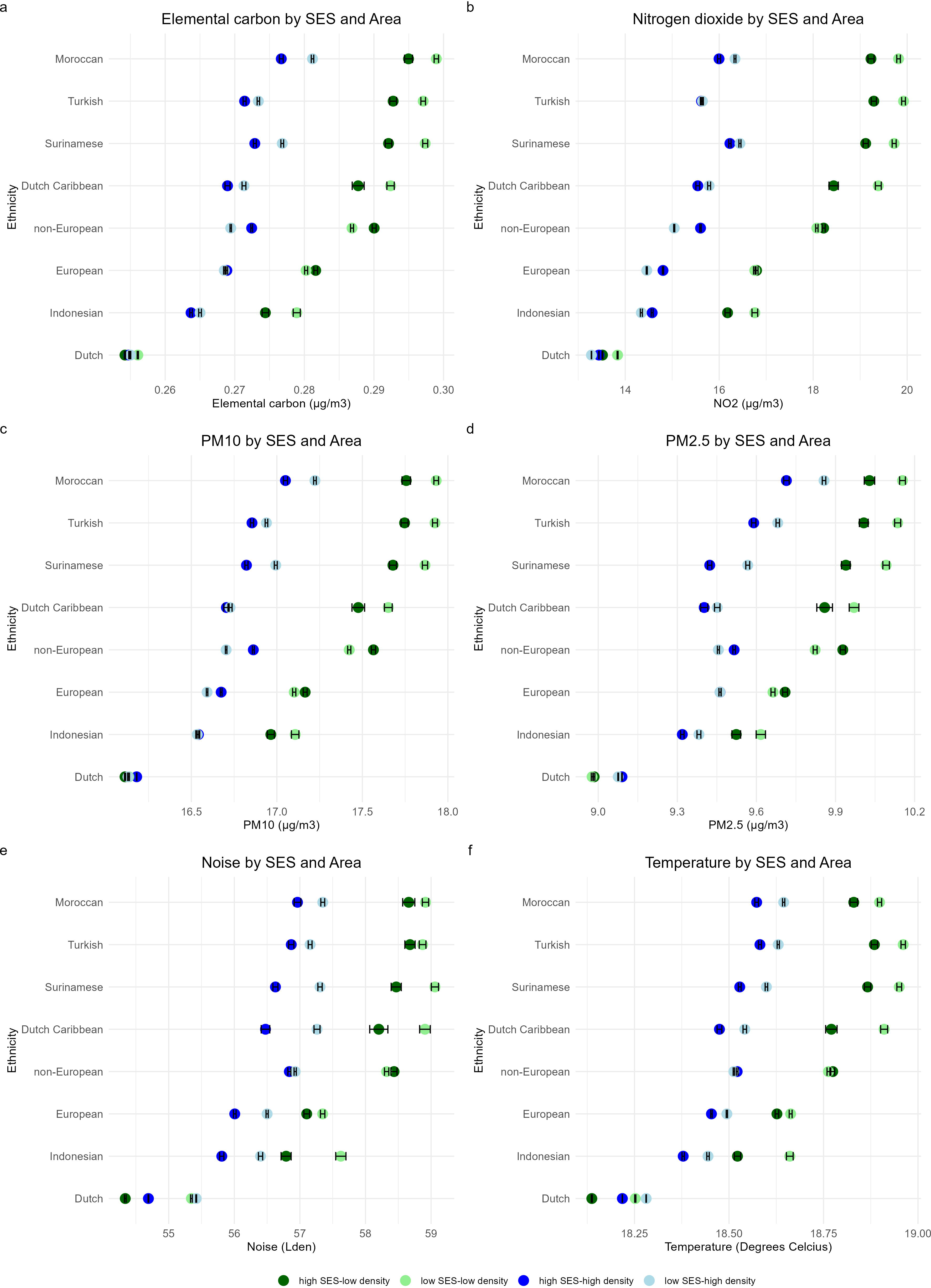
Estimated marginal means of physicochemical exposures among ethnic groups, by socio-economic status and population density.

### 3.3 Food and physical activity environment

The food and physical activity environments were generally less favorable for the Dutch-origin population compared to ethnic minority populations (Figure 2). For instance, The Food Environment Healthiness Index was 0.03 points higher [95%-CI 0.02; 0.03]) for Moroccans compared to Dutch-origin individuals in the “low SES-low population density” category. However, no noticeable ethnic differences were observed for the food environment in the “high SES-high population density” category. The physical activity environment was most favorable for Surinamese, Moroccans, Turks and Dutch Caribbeans, followed closely by non-Europeans and Europeans. For example, Dutch Caribbeans had a 1.98-point [95%-CI 1.92; 2.04] higher density of sport accommodations within a 1000m radius compared to Dutch-origin. Among the Dutch-origin population, small variations in estimated marginal means for walkability, bike path density, sport accommodation density and access to greenspace were observed by SES and population density. However, for ethnic minority populations, variations by population density were larger, which also increased the ethnic differences. Walkability, bike path density and sport accommodation density were higher in low than high population density areas, while access to greenspace was greater in high compared to low population density areas. Ethnic differences in bike path density between Turks and Dutch-origin individuals was 0.79 km/ha [95%-CI 0.78-0.79] in the “low SES-high population density” category compared to 1.82 km/ha [95%-CI 1.80; 1.83] in the “low SES-low population” density category.

**Fig 2:**
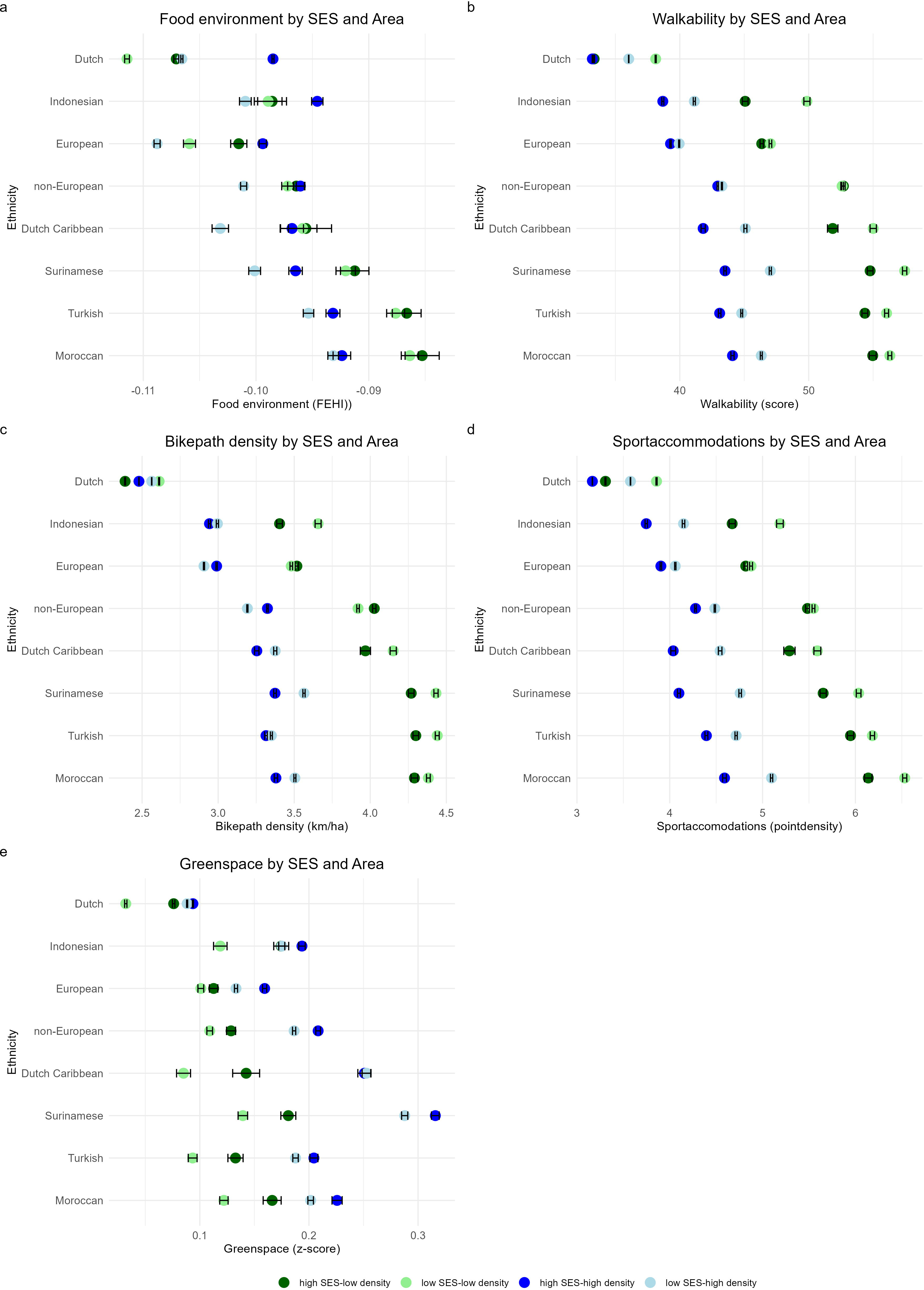
Estimated marginal means of food and physical activity environment amount ethnic groups, by socio-economic status and population density.

### 3.4 Socio-economic characteristics of the environment

Generally, the socio-economic characteristics of the environment were less favorable for ethnic minority compared to Dutch-origin individuals, with substantial differences between ethnic minority groups (Figure 3). Ethnic differences in liveability scores were generally small, especially for Indonesians, Europeans and non-Europeans compared to Dutch-origin. The largest differences were observed in the ‘high SES-low population density’ category for Moroccans compared to Dutch-origin: −0.19 points [95%-CI: −0.19; −0.19]. Area-level socio-economic position was highest among Dutch-origin, Indonesians and Europeans, and lowest for Moroccans and Turks. Ethnic differences were the smallest in the “high SES-high population density” categories. Notably, large ethnic differences in area-level socio-economic position were observed not only in the low SES categories, but also for the “high SES-low population” category among Moroccans, Turks, Surinamese and slightly for Dutch Caribbeans (e.g. −1.35 points [95%-CI −1.37; −1.34] for Turks compared to Dutch-origin). Similar patterns were observed for election turnout, living below the social minimum and labor participation. Ethnic differences were generally larger in low population density areas compared to high population density areas. For example, the difference in election turnout between Dutch Caribbeans and Dutch-origin was −4.23% [95%-CI −4.35; −4.11] in “high income-high population density” and −9.20% [95%-CI: −9.45; 8.96) in “high income-low population density” areas.

**Fig 3:**
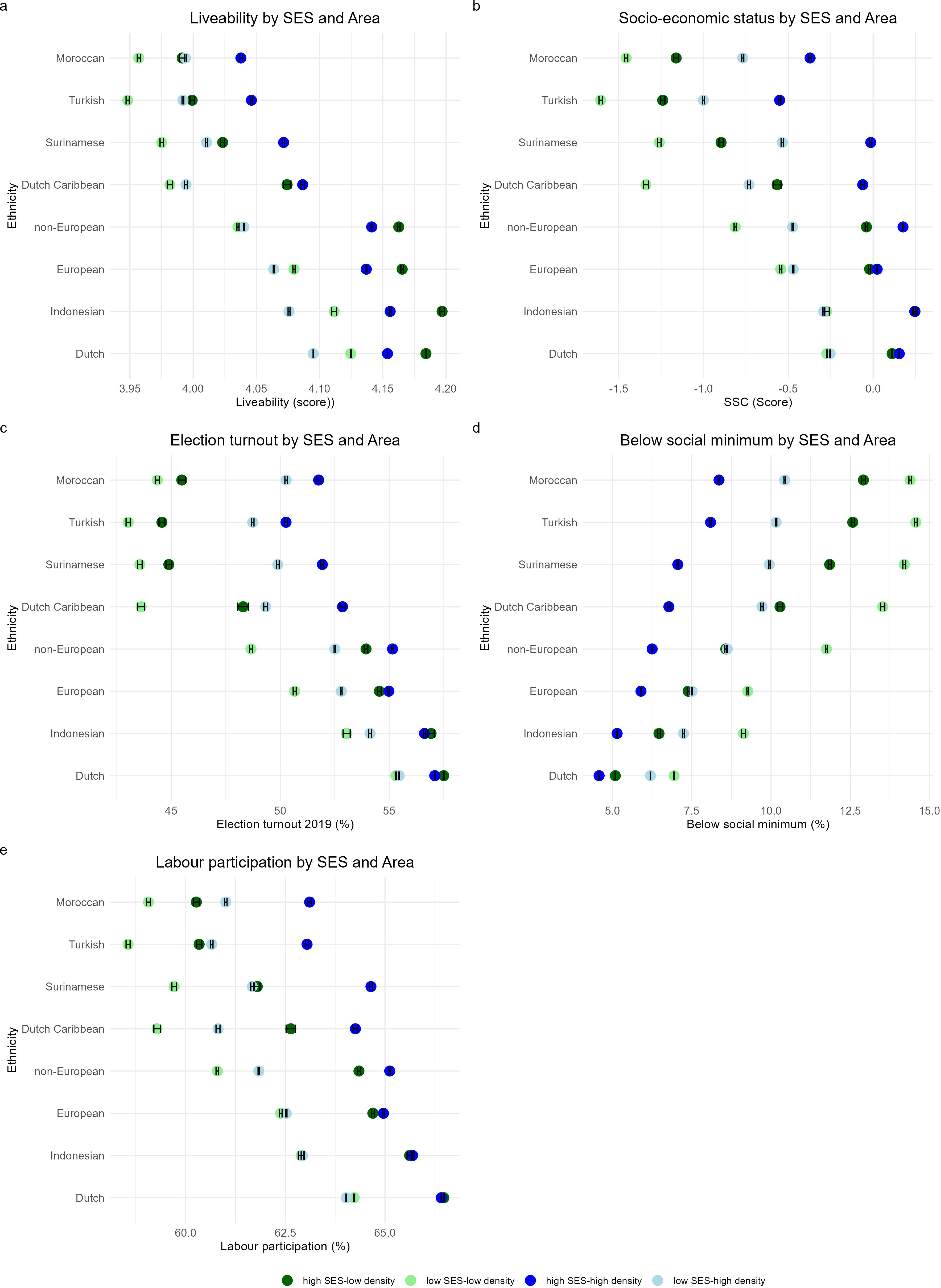
Estimated marginal means of socioeconomic characteristics of the environment among ethnic groups by socio-economic status and population density.

### 3.5 Health and social well-being in the environment

Ethnic differences in variables related to health and social well-being varied by specific variable (Figure 4). The prevalence of overweight at the neighborhood level was highest in “low SES-high population density” areas, with small ethnic differences. The estimated marginal mean ranged from 50.2% [95%-CI 50.1; 50.2] among Indonesians to 52.9% [95%-CI 52.8; 52.9] among Turks. Ethnic differences were largest in “high SES-low density” areas, where the prevalence of overweight was −4.15% [95%-CI −4.24; −4.06] among non-Europeans compared to Dutch-origin, and 2.92% [95%-CI 2.77;3.07] for Turks compared to Dutch-origin.

**Fig 4:**
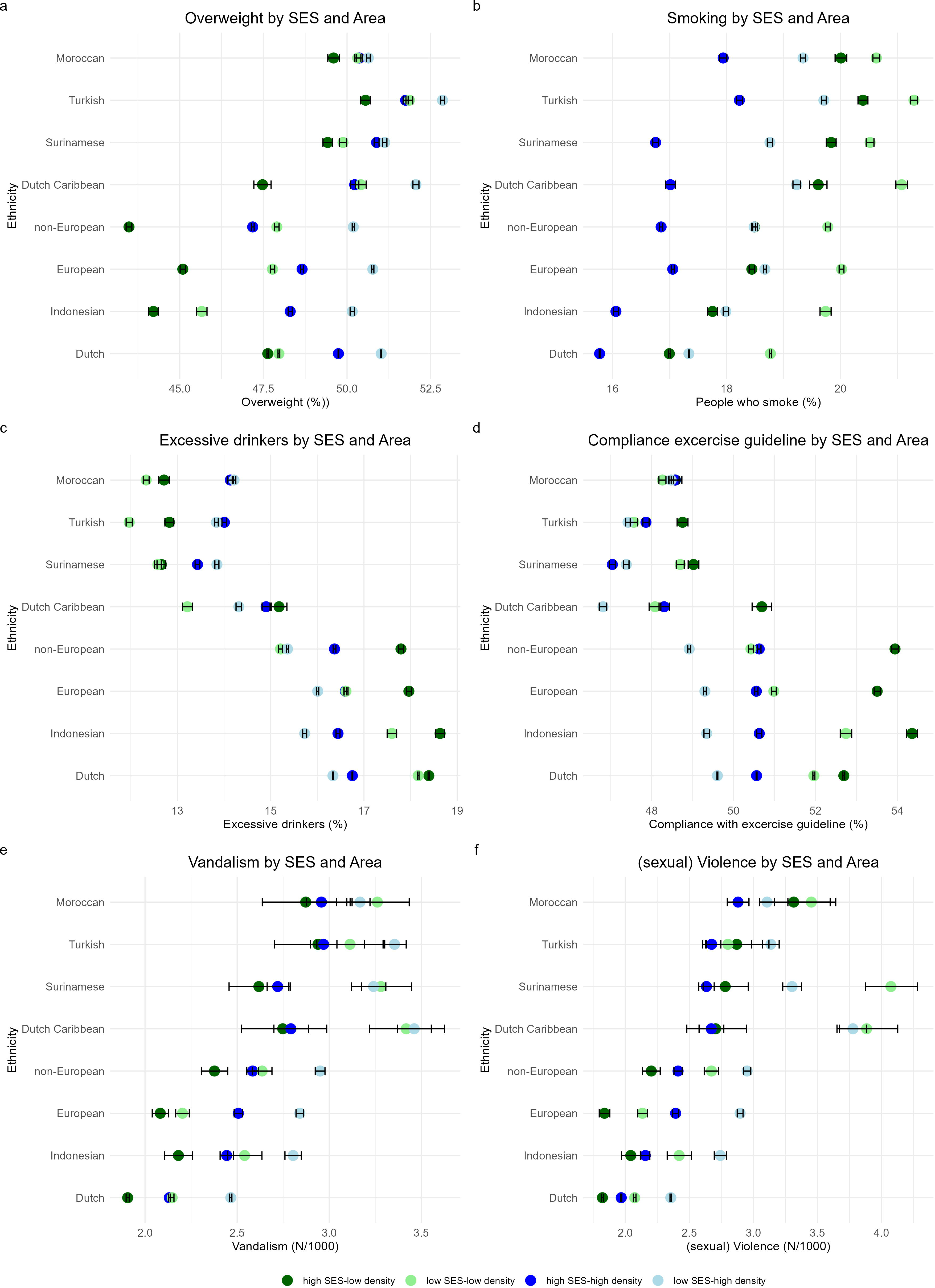
Estimated marginal means of health and social well-being in the environment by socio-economic status and population density.

The prevalence of smoking at the neighborhood level was lowest among Dutch-origin and highest among Turks and Moroccans (e.g. 2.52% [95%-CI 2.45; 2.58] for Turks compared to Dutch-origin). Prevalence of excessive drinking at the neighborhood level was highest among Dutch-origin, Indonesians, Europeans and non-European, and lowest among Moroccans, Turks and Surinamese. The ethnic differences were largest in low population density areas; for example, the difference for Surinamese compared to Dutch-origin was −5.74% [95%-CI −5.83; −5.64] in “high SES-low population density” areas. Compliance with exercise guidelines at the neighborhood level was lowest among Moroccans, Turks, Surinamese and Dutch Caribbeans. Compliance varied by population density among non-Europeans, Europeans, Indonesians and Dutch-origin, leading to larger ethnic differences in these areas. For example, compliance was −4.39% [95%-CI: −4.49; −4.30] around Turks compared to Dutch-origin. Finally, exposure to vandalism and (sexual) violence was lowest around Dutch-origin and highest among Dutch Caribbean, Moroccans, Turks and Surinamese. Ethnic differences in exposure were particularly large for Surinamese and Dutch Caribbeans in low SES areas (e.g. 0.50 per 1000 inhabitants per year [95%-CI 0.46; 0.54] for Surinamese compared to Dutch-origin in the “low SES-low population density” category.

### 3.6 Sensitivity and additional analyses

Ethnic differences in environmental exposures were generally slightly smaller for second-generation compared to first-generation migrants (Supplementary Figures 2-5). For instance, the differences in PM10 exposure was 1.90 µg/m^3^ [95%-CI: 1.88; 1.92] for first-generation Turks compared to Dutch-origin in the “low SES-low population density” category, but reduced to 1.46 µg/m^3^ [95%-CI 1.43; 1.48] for second-generation Turks. Similarly, first vs second generation Surinamese in the “high SES-low population density” category were exposed to 4.38 [95%-CI 4.28; 4.49] higher noise levels compared to Dutch-origin, while second-generation Surinamese had a higher noise exposure of 3.83 Lden [95%-CI 3.72; 3.95]. Most other exposures showed similar trends, even when Dutch-origin were the worst off, except for greenspace among Moroccans, Turks and non-Europeans. Notably, second-generation Indonesian migrants experienced better liveability and socio-economic position scores than Dutch-origin. No clear patterns were observed for overweight and compliance with exercise guidelines. While exposure to vandalism and (sexual) violence decreased slightly for most ethnic groups among second-generation migrants, this was not the case for Moroccans. Sensitivity analyses using different buffer zones or radii were yielded similar results (data not shown). Additionally, using different variables for socio-economic characteristics and health and social well-being of/in the environment generally reinforced our findings (Supplementary Figures 6 and 7). Finally, each ethnic group tended to have higher proportions, e.g. Moroccans, tended to have higher proportions of their own ethnic group living in their environment compared to Dutch-origin individuals, and this patterns was consistent across ethnic minority populations (Supplementary Figure 8).

## 4. Discussion

Physical and social health-related neighborhood exposures differed between ethnic groups living in the Netherlands, with the largest differences observed among Moroccans, Turks, Surinamese and Dutch Caribbeans compared to Dutch-origin. Physico-chemical exposures were higher among ethnic minority populations compared to Dutch-origin, while the food and physical activity environment was more favorable for these groups compared to Dutch-origin. Socio-economic characteristics of the environment were generally more positive for Dutch-origin. Ethnic differences in health and social well-being varied across factors, with safety factors being most favorable for Dutch-origin. These differences were more pronounced in low population density areas compared to high population density areas, and slightly smaller for high compared to low SES. Ethnic differences were persistent in second-generation migrants, although they were generally somewhat smaller compared to first-generation migrants.

### 4.1 Physico-chemical exposures

Ethnic minority populations experienced higher levels of physico-chemical exposures, such as air pollution, noise and temperature, compared to Dutch-origin. These exposures negatively impact physical and mental health, including all-cause mortality, cardiovascular- and respiratory diseases (Basner et al. 2014; Pope et al. 1995). The disproportionate exposure aligns with previous findings on ethnic differences in air pollution (Mustansar et al. 2025; van den Brekel et al. 2024), and social inequalities in environmental noise exposure (Dreger et al. 2019). Although less evidence is available regarding temperature, a review from the United States highlighted racial and socioeconomic disparities in heat-related health effects (Gronlund 2014), potentially linked to factors such as heat-absorbing surfaces and air conditioning ownership.

### 4.2 Food and physical activity environment

In contrast, the food and physical activity environment was more favorable among ethnic minority populations compared to Dutch-origin. This is in contrast with earlier studies of the food environment. A study in the United States showed Asian Americans and Blacks in non-metropolitan areas to be more likely to live in neighborhoods with lower access to healthy food options (Jiang et al. 2023). Moreover, a study in the Netherlands, limited to Amsterdam, showed that South-Asian Surinamese, African Surinamese, Turks and Moroccans were exposed to a less healthy food environment than individuals of Dutch-origin (Poelman et al. 2021). The study also indicated that the less healthy food environment did not contribute to ethnic differences in diet quality. However a systematic review from the United States suggests that convenience store access is associated with negative health outcomes in Hispanic and Black youth (Kraft et al. 2020).

The findings on the physical activity environment are consistent with a study from the United States showing higher walkability in neighborhoods with high proportions of non-Hispanic Blacks, Hispanics, Asians/Pacific Islanders/Native Americans compared to non-Hispanic white neighborhoods (King and Clarke 2015). While these factors are associated with health-promoting behaviors (James et al. 2015), their benefits depend on actual usage. A study on ethnic variations in park use and physical activity indicated park use may differ by ethnicity, necessitating additional efforts for ethnic minority populations to experience similar benefits (Derose et al. 2015). A review on urban park access found that although proximity to parks was slightly better for ethnic minority populations, park acreage and quality were worse (Rigolon 2016).

### 4.3 Socio-economic characteristics of the environment

Socio-economic characteristics of the environment were generally more favorable for Dutch-origin compared to ethnic minority populations, as reflected in higher liveability scores, greater election turnout, higher labor participation, and lower rates of living below the social minimum. While no studies are available specifically at the neighborhood level, similar findings have been observed in studies focusing on individuals (Nazroo et al. 2020). Moreover, our results align with the observation that ethnic minority populations often reside in more deprived areas (Ng Fat et al. 2023). These disparities highlight structural barriers that may hinder ethnic minority populations from accessing resources and opportunities for social mobility and well-being.

### 4.4 Health and social well-being in the environment

Differing patterns in health and social well-being at the neighborhood level were observed. Prevalence of excessive drinking at the neighborhood level was most prevalent for Dutch-origin, while prevalence of smoking at the neighborhood level was more common among ethnic minority populations. These health-related behaviors within communities can influence individual health by social contagion (Powell et al. 2015). However, social norms from friends and family may play a more significant role than those of neighbors (Dulin et al. 2018), potentially affecting health behaviors in ways not fully captured by home addresses alone. Exposure to vandalism and (sexual) violence was most favorable for Dutch-origin and less so for ethnic minority populations. Perceptions of safety and actual safety levels are critical determinants of both mental health and quality of life (Lorenc et al. 2012). Inequalities may stem from structural inequalities, including differences in neighborhood investments, policing practices, and historical segregation patterns.

### 4.5 Urban vs. rural differences

Interestingly, ethnic inequalities in physical and social neighborhood exposures environmental factors were generally more pronounced in low population density compared to high population density areas, likely due to smaller variations in exposures in high population density areas. This finding challenges the narrative that urban environments exacerbate inequalities (Sarkar et al. 2024) and suggests that rural settings may pose unique challenges for ethnic minority populations. Rural areas often have fewer healthcare resources, lower socioeconomic opportunities, and less robust public infrastructure which can disproportionally impact ethnic minority populations (Swope and Hernández 2019). Future studies should delve deeper into these rural-urban disparities to inform tailored policy interventions and reduce inequities in both settings.

### 4.6 Strengths and limitations

Our study included a wide range of both physical and social neighborhood exposures, providing a broad overview of ethnic inequalities. A strength is the use of individual-level data for all adult Dutch inhabitants, with minimal missing data. Additionally, our study went beyond air pollution, including many underexplored environmental factors, addressing a critical gap in the literature. However, there are limitations. Using home address as a proxy for environmental exposure may not fully capture the complexity of factors influencing health outcomes. For example, noise and temperature exposures may be moderated by home characteristics, such as building quality or air conditioning presence. The food environment may be more relevant in workplaces or during commuting rather than solely around the home (Burgoine and Monsivais 2013). Additionally, asylum seekers might be somewhat underrepresented in our study population due to registration delays of up to six months after arrival in the Netherlands.

### 4.7 Conclusions

This study highlight the complex associations between ethnicity and physical and social health-related neighborhood exposures in the Netherlands. Physico-chemical exposure, socio-economic characteristics of the environment and environmental safety were less favorable among ethnic minority populations compared to the Dutch-origin population. The food and physical activity environment was more favorable for most ethnic minority populations. Ethnic inequalities were most pronounced among Moroccans, Turks, Surinamese and Dutch Caribbeans compared to Dutch-origin. These findings contribute to the broader literature on environmental determinants of health and equity and call for further research to uncover the mechanisms driving these inequalities and evaluate the effectiveness of interventions designed to promote equity and well-being across diverse populations. This underscores the urgent need for targeted interventions to mitigate health inequities among ethnic minority populations.

## Data statement

We were granted access to the data, but are not the owners of the data. Data can be requested for statistical and scientific research from Statistics Netherlands by contacting. Data from the health monitor is freely available through: StatLine - Gezondheid per wijk en buurt; 2012/2016/2020/2022 (indeling 2022). Data from the Geoscience and Health Cohort Consortium can be requested through www.gecco.nl.

## Author contributions

Conceptualization: MM, FR, JL, KS, JB; Formal analysis: MM; Funding acquisition: MM, JB; Methodology: MM, FR, JL, KS, JB; Project administration: MM, Supervision: JB; Visualization: MM; Writing - original draft: MM; Writing – review and editing: MM, FR, JL, PE, MB, KS, IV, JB.

## Data Availability

https://data.overheid.nl/en/dataset/42936-gezondheid-per-wijk-en-buurt--2012-2016-2020-2022--indeling-2022-

https://www.gecco.nl/

https://www.cbs.nl/nl-nl/onze-diensten/maatwerk-en-microdata/microdata-zelf-onderzoek-doen

