## Supplementary for "Ethnic inequalities in physico-chemical, physical and social neighborhood exposures: An individual-level data analysis of 13,926,781 adults"

**Supplementary Figure 1: Flow-diagram of included study population**

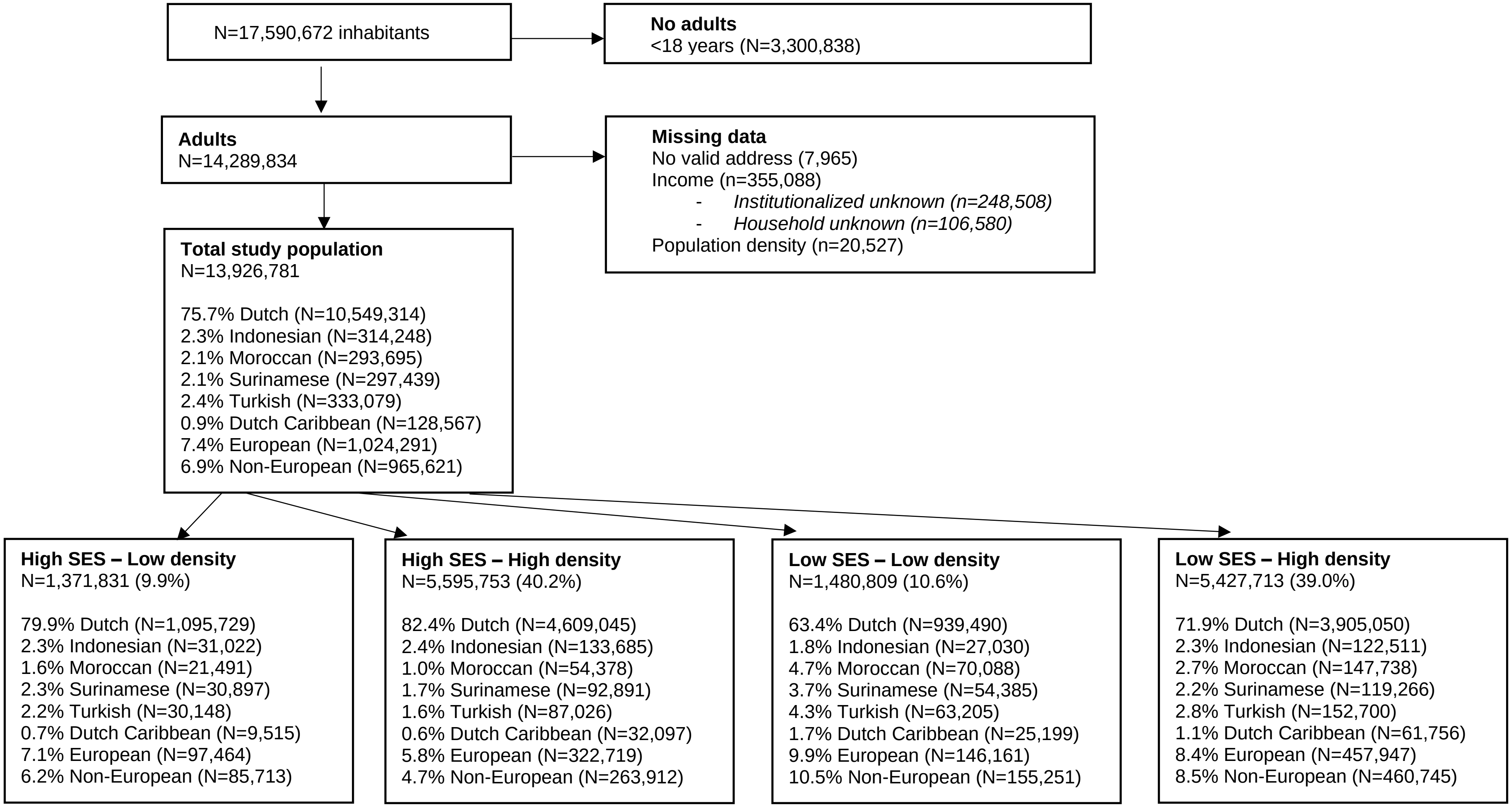

**Supplementary Figure 2: Estimated marginal means of physicochemical exposures among different ethnic groups considering migration generation, stratified by socio-economic status and population density**

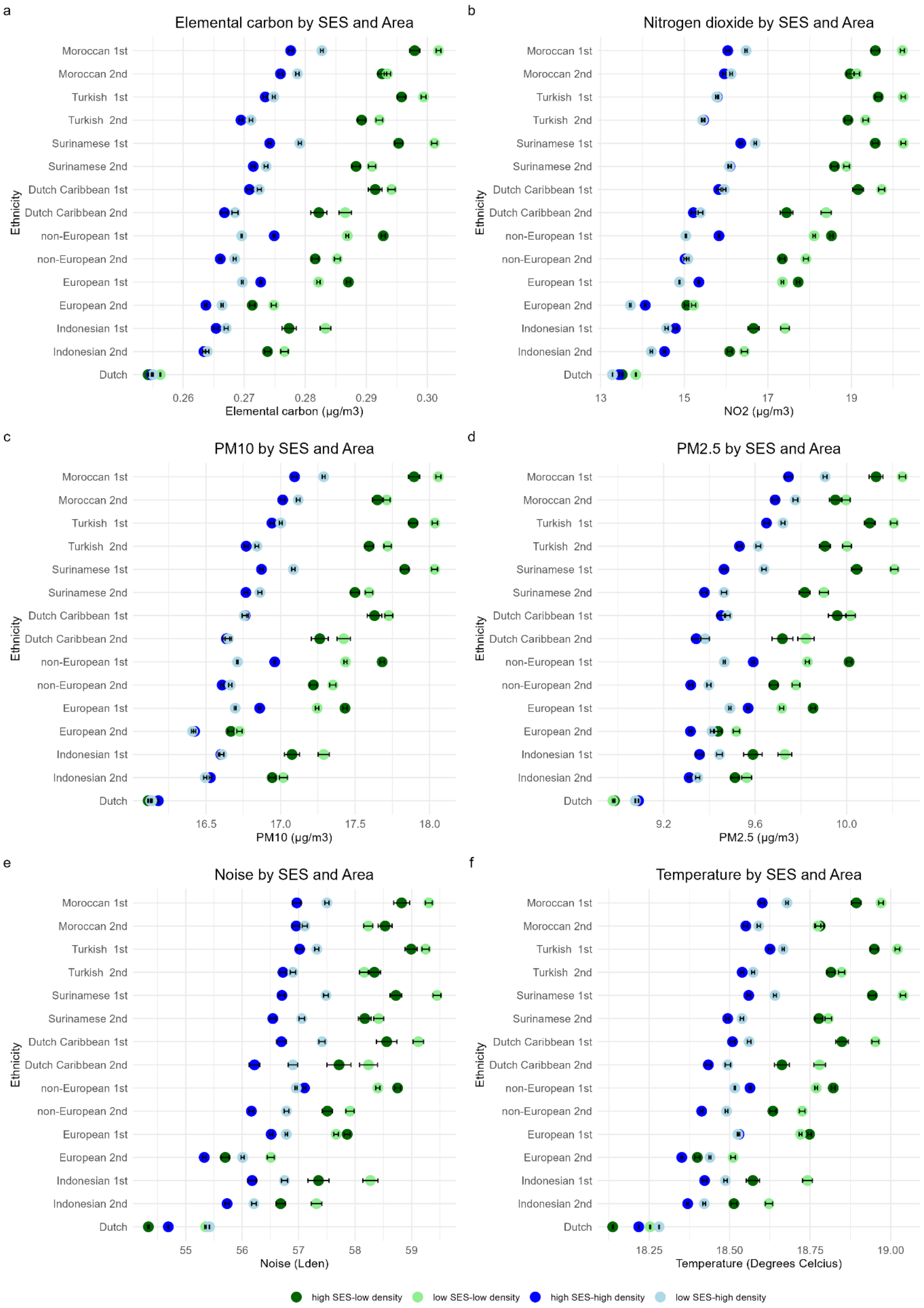

**Supplementary Figure 3: Estimated marginal means of the food and physical activity environment among different ethnic groups, stratified by socio-economic status and population density**

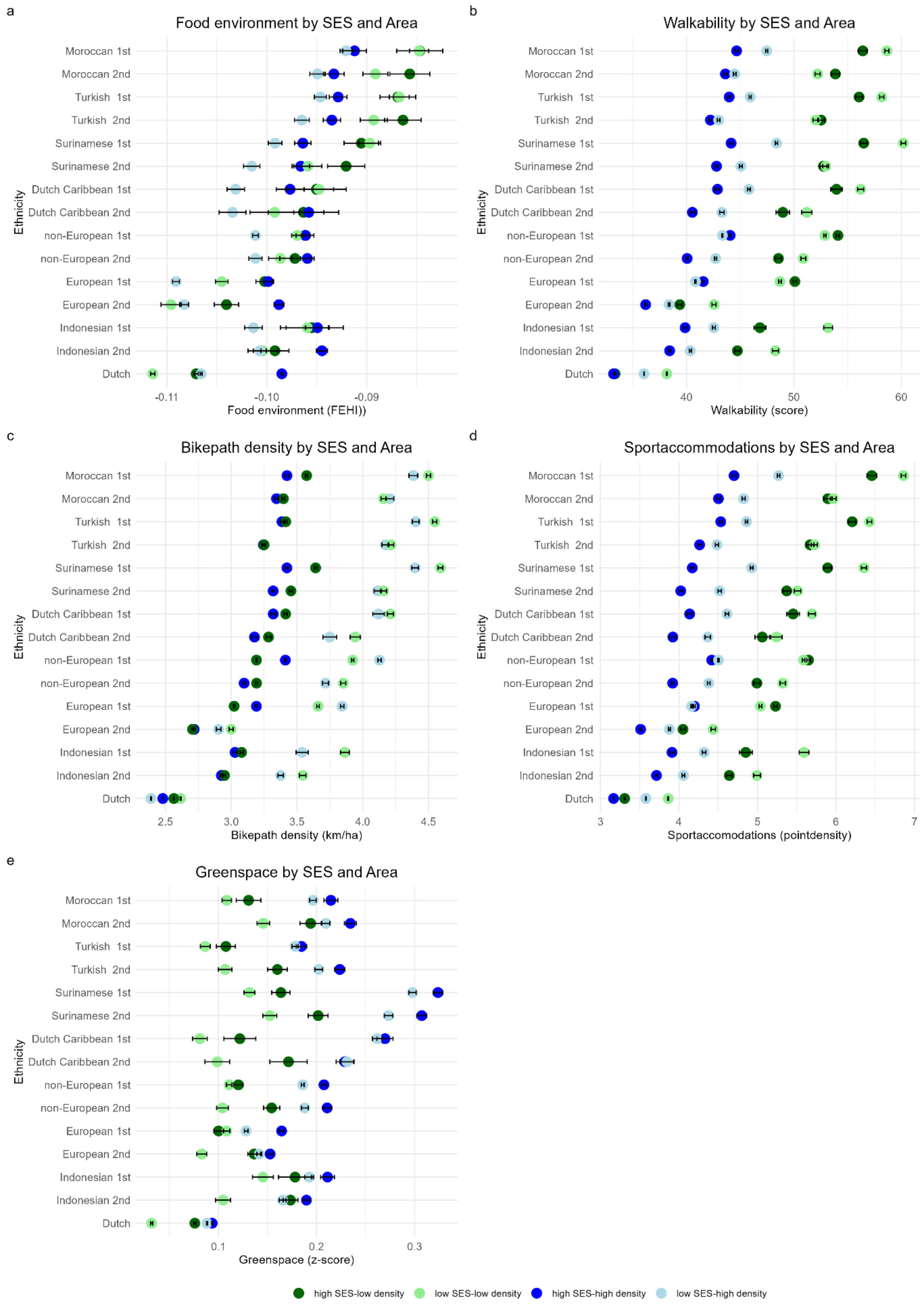

**Supplementary Figure 4: Estimated marginal means of socioeconomic characteristics of the environment among different ethnic groups, stratified by socio-economic status and population density**

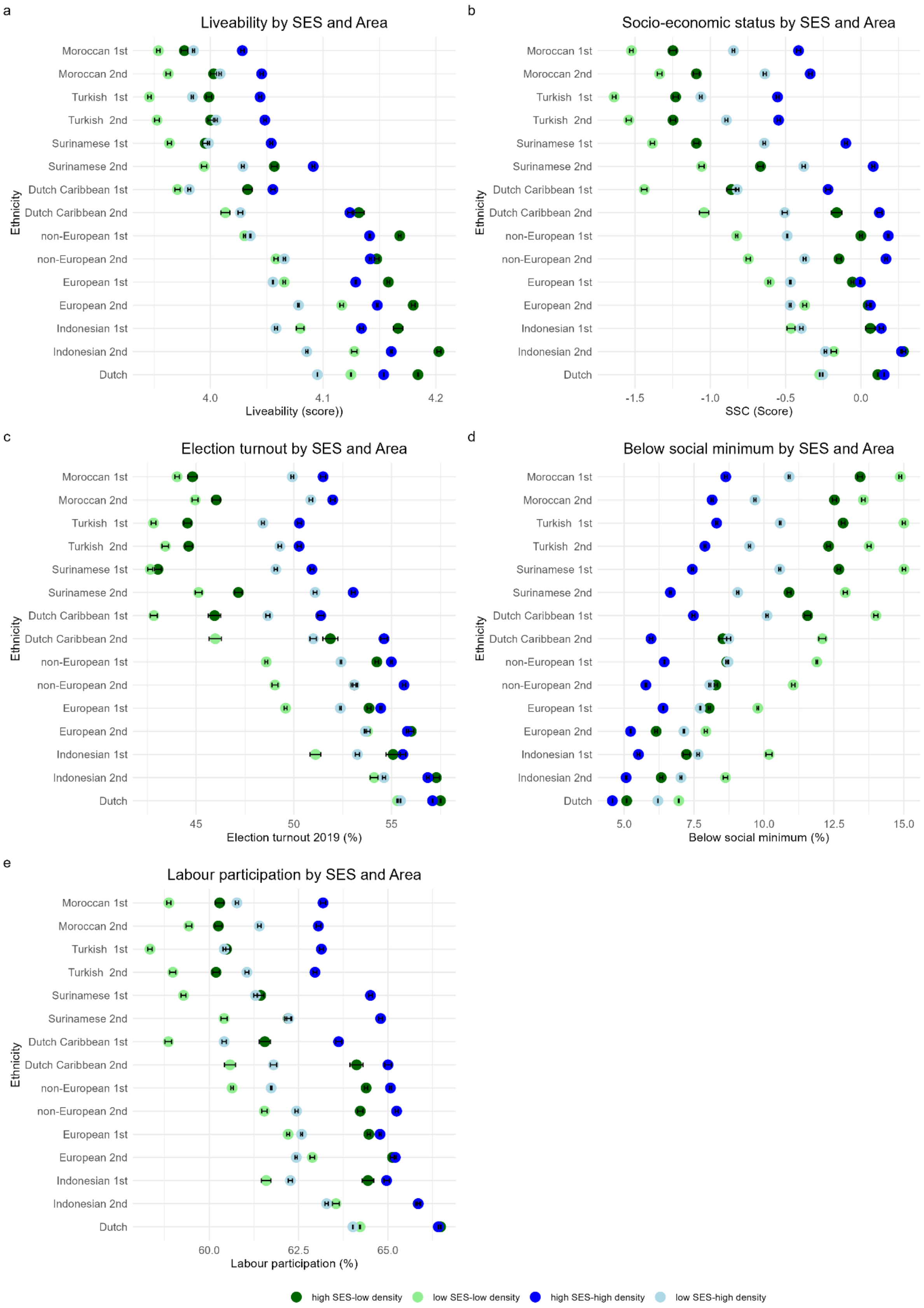

**Supplementary Figure 5: Estimated marginal means of health and social well-being in the environment among different ethnic groups, stratified by socio-economic status and population density**

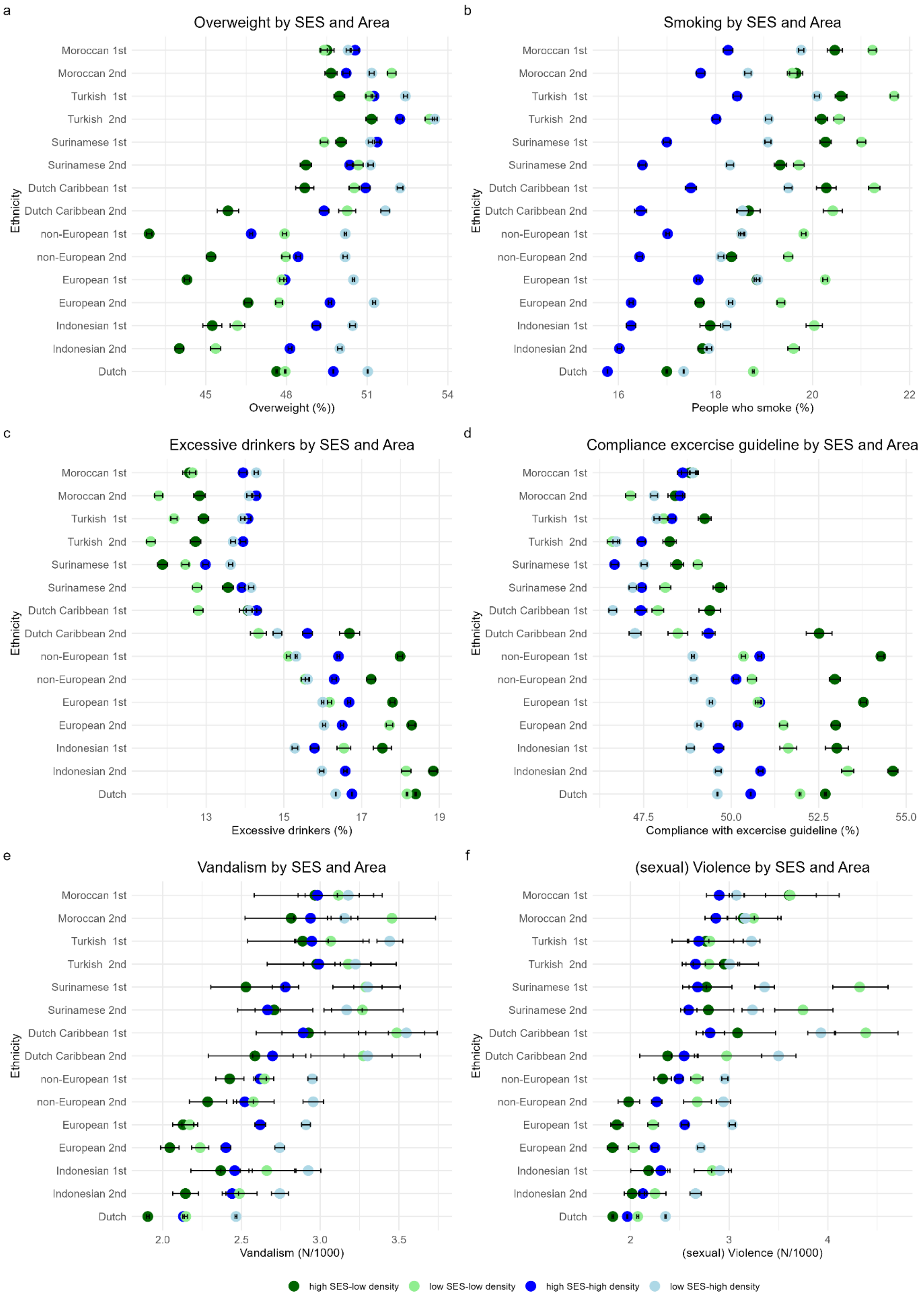

**Supplementary Figure 6: Estimated marginal means of additional socioeconomic characteristics of the environment among different ethnic groups considering migration generation, stratified by socio-economic status and population density.**

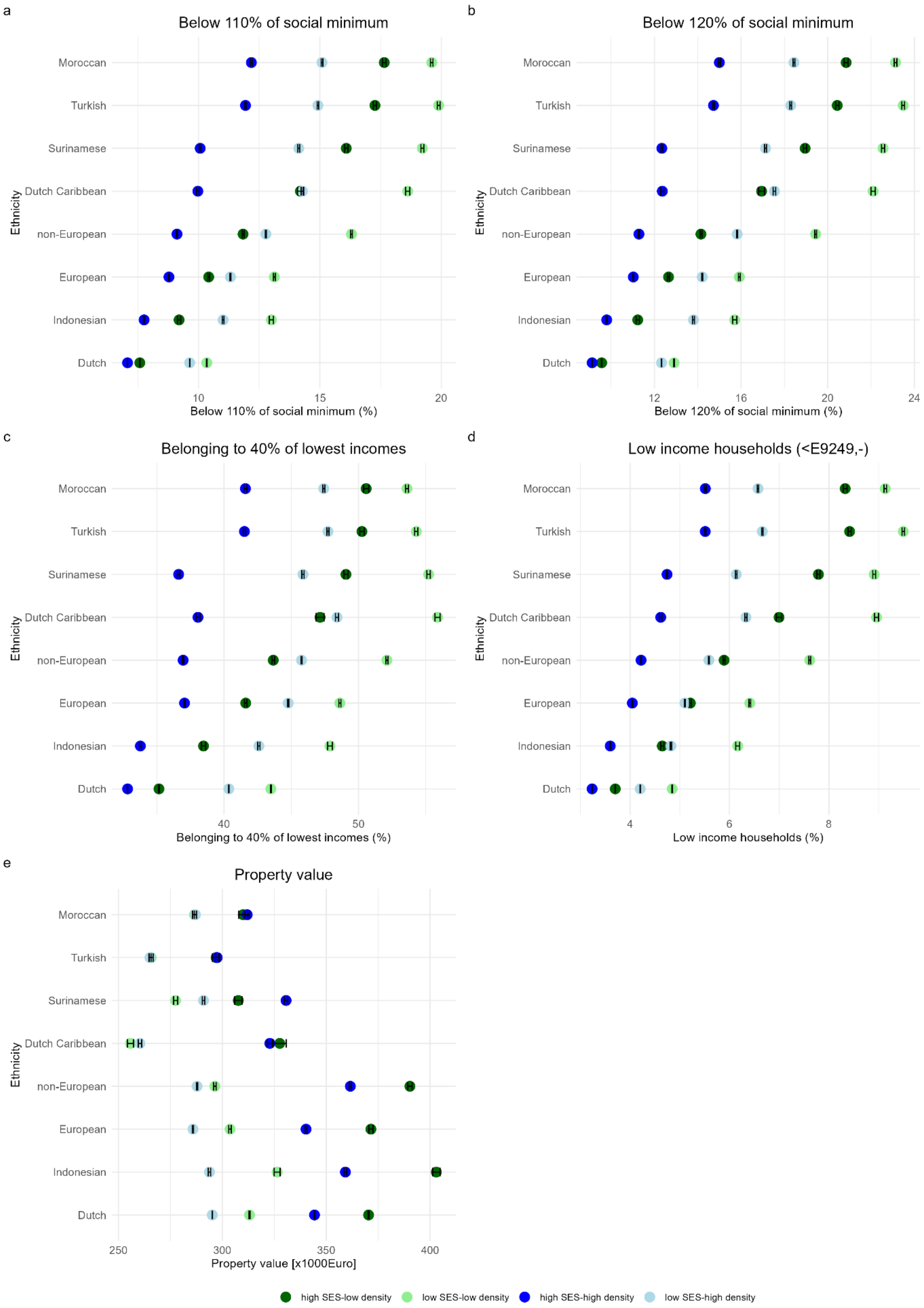

**Supplementary Figure 7: Estimated marginal means of health and social well-being in the environment among different ethnic groups considering migration generation, stratified by socio-economic status and population density**

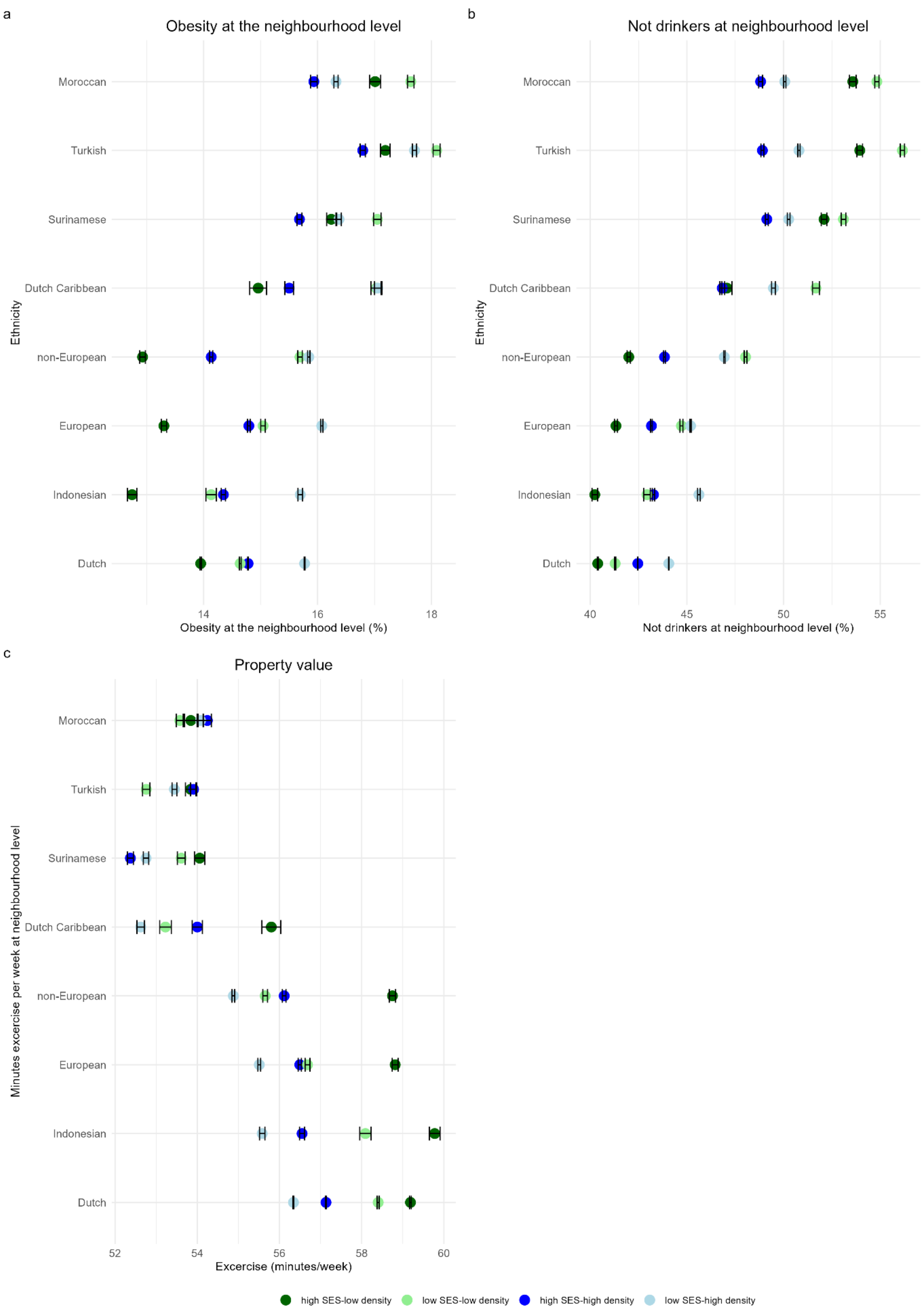

**Supplementary Figure 8: Estimated marginal mean percentages of Moroccan, Surinamese, and Dutch Caribbean people in the neighbourhood of different ethnicities considering migration generation.**

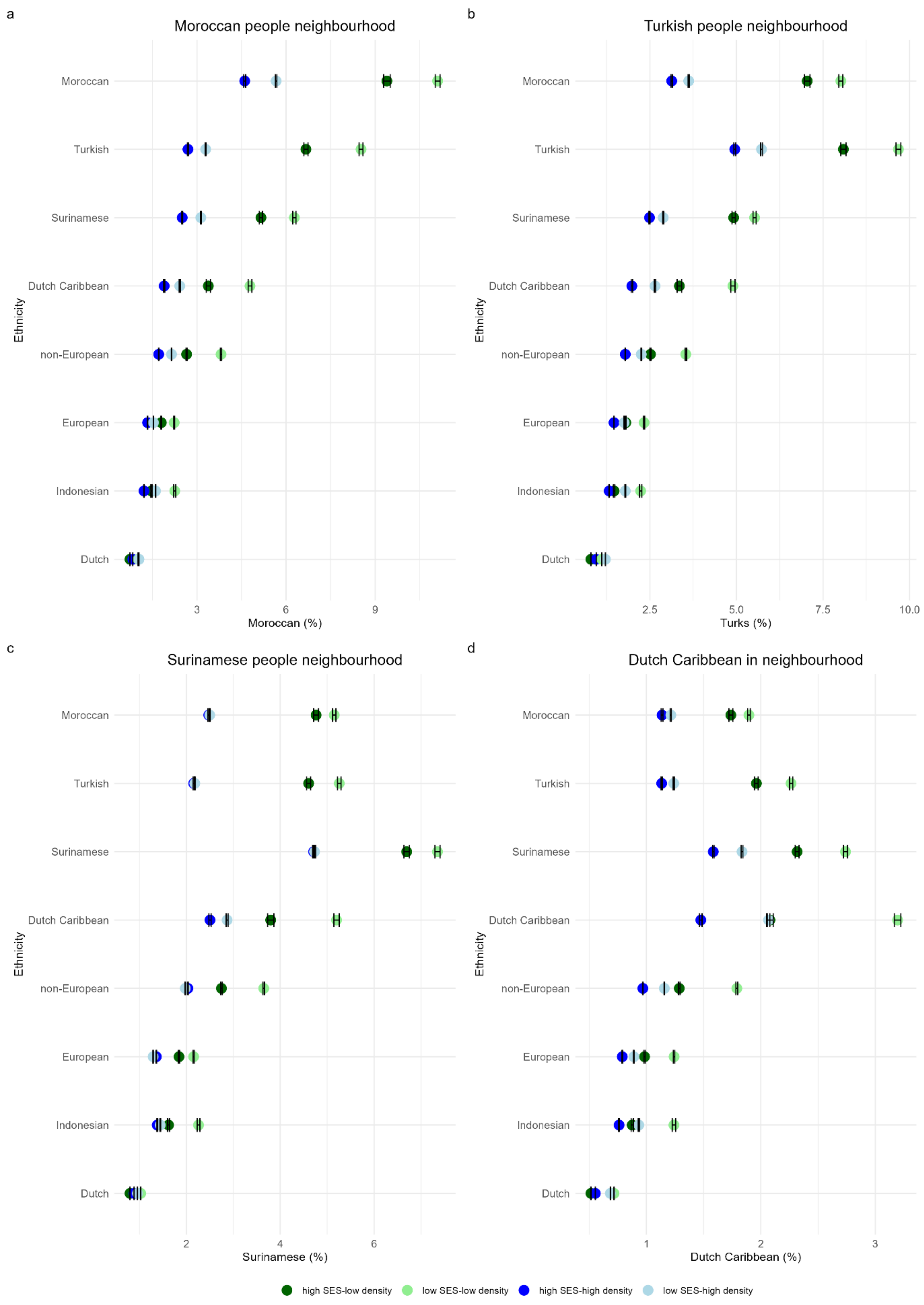
